# Is OR “portable” in meta-analysis? Time to consider bivariate generalized linear mixed model

**DOI:** 10.1101/2020.11.05.20226811

**Authors:** Mengli Xiao, Yong Chen, Stephen Cole, Richard MacLehose, David Richardson, Haitao Chu

**Affiliations:** Division of Biostatistics, School of Public Health, University of Minnesota, Minneapolis, MN 55455, USA; Department of Biostatistics and Epidemiology, University of Pennsylvania, Philadelphia, Pennsylvania 19104, USA; Department of Epidemiology, University of North Carolina, Chapel Hill NC 27599, USA; Division of Epidemiology and Community Health, School of Public Health, University of Minnesota, Minneapolis, MN 55455, USA

**Keywords:** baseline risk, Cochrane Database of Systematic Reviews (CDSR), correlation, meta-analysis, odds ratio, relative risks

## Abstract

**Objectives:** A recent paper by Doi et al. advocated completely replacing the relative risk (RR) with the odds ratio (OR) as the effect measure used to report the association between a treatment and a binary outcome in clinical trials and meta-analyses. Besides some practical advantages of RR over OR and the well-known issue of the OR being non-collapsible, Doi et al.’s key assumption that the OR is “portable” in the meta-analysis, i.e., study-specific ORs are likely not correlated with baseline risks, was not well justified.

**Study designs and settings:** We summarized the Spearman’s rank correlation coefficient between study-specific OR and the baseline risk in 40,243 meta-analyses from the Cochrane Database of Systematic Reviews (CDSR).

**Results:** Study-specific ORs are negatively correlated with baseline risk of disease (i.e., higher ORs tend to be observed in studies with lower baseline risks of disease) for most meta-analyses in CDSR. Using a meta-analysis comparing the effect of oral sumatriptan (100 mg) versus placebo on mitigating the acute headache at 2 hours after drug administration, we demonstrate that there is a strong negative correlation between OR (RR or RD) with the baseline risk and the conditional effects notably vary with baseline risks.

**Conclusions:** Replacing RR or RD with OR is currently unadvisable in clinical trials and meta-analyses. It is possible that no effect measure is “portable” in a meta-analysis. In cases where portability of the effect measure is challenging to satisfy, we suggest presenting the conditional effect based on the baseline risk using a bivariate generalized linear mixed model. The bivariate generalized linear mixed model can be used to account for correlation between the effect measure and baseline disease risk. Furthermore, in addition to the overall (or marginal) effect, we recommend that investigators also report the effects conditioning on the baseline risk.

**What is New?:** *Key findings:*

- In most meta-analyses in Cochrane Database of Systematic Reviews, there is notable negative correlation between ORs and baseline risks.
- When such a correlation is not negligible, the OR is not “portable” across studies with different baseline risks.
- When an effect measure is not “portable”, one may derive the effects conditioning on the baseline risk from a bivariate generalized linear mixed model.

*What this study adds to what was known:*

- The recommendation to replace the RR with the OR in clinical trials and meta-analyses is misguided.
- The OR is not a better effect summary than RR and RD in a single study or in meta-analyses; the noncollapsibility of OR can lead to misleading results in a single study and the OR is generally not portable in the meta-analysis.
- In addition to reporting effect measures such as the OR, RR or RD, it is also important to present the baseline risk.

*What is the implication and what should change now?:*

- When none of the effects are “portable” in a meta-analysis, in addition to report the overall (or marginal) effect, one should also report the effects conditioning on the baseline risk, regardless of the measure of choice.

## 1. Introduction

We congratulate Doi et al.^1^ for their thought-provoking, yet controversial, paper that advocates for the use of the odds ratio (OR) over relative risk (RR) as a measure of effect. They suggest that clinical trials and meta-analyses should use the OR instead of RR as the effect measure for binary outcomes. Doi et al. demonstrate that the magnitude of the RR varies with outcome prevalence. They reached such a conclusion by varying the prevalence of a hypothetical outcome and comparing RRs, while assuming a fixed OR that was treated as the true measure of associations due to its independence with prevalence, (i.e., portability). Although such a recommendation was supported by technically rigorous mathematical derivations, we believe that the call to end the primary use of RR in clinical trials and meta-analyses and to replace the RR by the OR is misguided.

ORs are notoriously difficult to interpret. When people hear “odds” they think of “risks” and this leads to the common misinterpretation of the OR as a RR by scientists and the public, which is a serious concern.^2-7^ For example, an OR of 2 is not generally a doubling of risk (if the risk in the control group is 20% and the OR is 2, then the risk in the treated group is 33.3% not 40%). In contrast, the RD and RR offer clearer interpretations. A RD of 0.1 means that the risk of the outcome in the treated group is simply a 10 percentage points higher than the risk of the outcome in the control groups. A RR of 2 means that the risk is doubled in the treatment group compared to the control group. Such a distinction in ease of description has motivated efforts to transform ORs to RRs, or use various regression models to estimate the RR or RD directly.^3,8-12^ If RR was fully replaced by OR as Doi et al. recommend, spurious conclusions might become more common because the OR seems to be inevitably interpreted as a RR, and often this will lead people to overestimate the relative effect of a treatment.^2,7,8,13-15^ For example, when the risk is 0.8 in the treatment group and 0.4 in the control group, the corresponding OR=6 and RR=2. Because the OR = 6 will commonly be misinterpreted as an RR = 6 by the press, clinicians, and other researchers, such an exaggerated treatment effect can be misleading to evidence-based practices. ^2,5,7,14,15^

Although Doi et al argue that the variability of the RR (and, by extension, the RD) with baseline prevalence is an impediment to using those measures, we argue the opposite: that the variability of the RR with baseline prevalence is in fact an *appealing* feature in the utility of these measures. The risk of the outcome in the control group affects the clinical meaning of effect measures, as it should.^4,16,17^ For example, when the baseline risk is extremely low, say 0.000,001, and the risk after treatment is 0.000,01, the OR and RR, which are both about 10, do not show how rare the outcome is; on the other hand, the RD is 0.000,009 and conveys the small increase in risk. If the baseline risk is high, say 0.96, and the risk is 0.995 for the treatment group, a small effect is expected, which can be reflected by RR and RD (RR=1.04, RD=0.035) rather than the corresponding OR (8.29). Because the OR can drastically overstate the magnitude of effect when the outcome is common and is approximately the same as the RR if the outcome is rare, we do not see a general need for the OR as a default measure of effect. In summary, the interpretation of an effect measure should account for the magnitude of the baseline risk.

In addition to the potential misleading interpretation of the *magnitude* of OR as an effect size measure, numerous studies have discussed the fact that the OR lacks an important and appealing property for interpreting effect estimates, i.e., collapsibility.^18-28^ In a single study with a non-confounding stratification variable, if the stratum-specific effects are homogenous, then they are expected to be the same as the crude effect, a desirable property known as collapsibility of an effect measure. Both RR and RD have this desirable property, while OR does not.^19,24,29^ We will discuss this noncollapsibility issue of OR in details in Section 2.

Furthermore, Doi et al. argued that the OR is a better effect measure of association because OR is likely unassociated with the baseline risk. We believe that a careful discussion is needed about the practicality of, and empirical support for, this assumption. In meta-analyses, differences in the baseline risk across studies may explain the treatment effect heterogeneity.^30-32^ Multiple studies suggest that understanding the treatment effect without considering potential variation in treatment effects between groups defined by factors associated with differences in baseline disease risk, is potentially misleading.^31-37^ Although Doi et al. showed that the OR was independent of the prevalence by pooling 140,620 trials across 14,960 meta-analyses altogether, it seems counterintuitive that OR is not related to the baseline risk; the analysis did not consider differences in meta-analyses, i.e. stratification by each meta-analysis. Hence, it remains unclear whether a general statement that OR better reflects the effect of interest can be made.

In this paper, we first expand in detail about how noncollapsibility makes OR an undesirable summary measure in Section 2. Then, we evaluate whether the OR is correlated with the baseline risk in meta-analysis in Section 3, i.e., whether OR is portable after the differences between meta-analyses are removed. Specifically, to investigate if the correlation commonly exists, we estimate the Spearman’s rank correlation coefficient for each meta-analysis with a large-scale evaluation of the meta-analyses from the Cochrane’s Database of Systematic Reviews (CDSR). Then in Section 4, when there is a correlation between the baseline risk and the effect measure (e.g. RR, RD or OR) within a meta-analysis, we recommend a bivariate generalized linear mixed-effects model (BGLMM). A detailed case study is presented to illustrate its implementation.

## 2. Is OR a better summary effect measure than RD and RR in a single study?

Unlike the RD or RR, the OR does not correspond to the proportional changes in average odds in the absence of confounding factors, which is known as the noncollapsibility.^18-21,27,29,38^ Consequently, the erroneous assumption that the crude and stratum-specific ORs will be equivalent in the absence of confounding can lead to substantial confusion. Specifically, when the ORs are homogenous across strata, the crude OR can be different from stratum-specific ORs in the absence of confounding. This is not true of the RD or RR which are collapsible in the absence of confounding. Table 1A and Table 1B, adapted from Greenland et al (1999), are examples that demonstrate the collapsibility of RD and RR in contrast to OR’s noncollapsibility across the covariate Z. It is important to note that there is no association between Z and X in these tables: P(Z=1|X=1) = P(Z=1|X=0) = 0.5. Because there is no association between Z and X, we know that Z is not a confounder. In Table 1A, we see that the RD is collapsible in the absence of confounding because its crude value is equal to the constant stratum-specific value, all of which are RD=0.2. This is not true for the OR. The stratum specific ORs are both 2.67 and, despite the fact that there is no confounding, the crude OR is 2.25. Although both the stratum-specific OR and crude OR are correctly calculated, one could falsely assume there is confounding bias from OR from Table 1A. Table 1B further demonstrates that RR can be collapsible; its crude value is equal to stratum-specific values in the absence of confounding.

**Table 1:** An illustration example to demonstrate the collapsibility of the RD and RR, and noncollapsibility of OR between outcome (Y), treatment (X) and strata (Z).

**A. The crude RD equals to a constant stratum-specific RD value**
|  | Z=1 |  | Z=0 |  | Crude |  |
| --- | --- | --- | --- | --- | --- | --- |
|  | X=1 | X=0 | X=1 | X=0 | X=1 | X=0 |
| Y=1 | 80 | 60 | 40 | 20 | 120 | 80 |
| Y=0 | 20 | 40 | 60 | 80 | 80 | 120 |
| Risk | 0.80 | 0.60 | 0.40 | 0.20 | 0.60 | 0.40 |
| RD | 0.20 |  | 0.20 |  | 0.20 |  |
| RR | 1.33 |  | 2.00 |  | 1.50* |  |
| OR | 2.67 |  | 2.67 |  | 2.25 |  |

\*, RR varies across the two strata, but it can still be collapsible across Z as it can be computed as the ratio of weighted averaged risks, i.e.,

**B. The crude RR equals to a constant stratum-specific RR value**
|  | Z=1 |  | Z=0 |  | Crude |  |
| --- | --- | --- | --- | --- | --- | --- |
|  | X=1 | X=0 | X=1 | X=0 | X=1 | X=0 |
| Y=1 | 60 | 30 | 40 | 20 | 100 | 50 |
| Y=0 | 40 | 70 | 60 | 80 | 100 | 150 |
| Risk | 0.60 | 0.30 | 0.40 | 0.20 | 0.50 | 0.25 |
| RD | 0.30 |  | 0.20 |  | 0.25* |  |
| RR | 2.00 |  | 2.00 |  | 2.00 |  |
| OR | 3.50 |  | 2.67 |  | 3.00 |  |

\*, RD can still be collapsible from different stratum-specific values in Table 1B by being the difference of weighted averaged risks, i.e.,
$$RD_{crude} = \text{averaged risk}_{X=1} - \text{averaged risk}_{X=0} = (\text{risk}_{X=1,Z=1}P(Z=1|X=1) + \text{risk}_{X=1,Z=0}P(Z=0|X=1)) - (\text{risk}_{X=0,Z=1}P(Z=1|X=0) + \text{risk}_{X=0,Z=0}P(Z=0|X=0)) = (0.6 \cdot 0.5 + 0.4 \cdot 0.5) - (0.3 \cdot 0.5 + 0.2 \cdot 0.5) = 0.25$$ On the contrary, OR is still not collapsible across Z because
$$\frac{\text{averaged odds}_{X=1}}{\text{averaged odds}_{X=0}} = \frac{\text{odds}_{X=1,Z=1}P(Z=1|X=1) + \text{odds}_{X=1,Z=0}P(Z=0|X=1)}{\text{odds}_{X=0,Z=1}P(Z=1|X=0) + \text{odds}_{X=0,Z=0}P(Z=0|X=0)} = \frac{\frac{0.6}{1-0.6} \cdot 0.5 + \frac{0.4}{1-0.4} \cdot 0.5}{\frac{0.3}{1-0.3} \cdot 0.5 + \frac{0.2}{1-0.2} \cdot 0.5} = 3.19 \neq OR_{marginal} = 3.0$$ RD: risk difference; RR: relative risks; OR: odds ratio.

Such noncollapsibility of OR may undermine the interpretability of a summary measure of association from a randomized controlled trials (RCTs) when considering the treatment effect within a subgroup of the treated. If potential confounders are well-controlled for by randomization, covariate adjustment is generally not necessary; and, often it is desirable to be able to interpret the crude treatment effect estimate as the stratum-specific effects.^39^ In a review of 288 RCT with binary outcomes, 12.4% of them evaluated the primary outcome by the crude OR.^40^ One may interpret these crude ORs as the amount the odds of the outcome would change under the intervention in the whole population; however, that same OR does not apply to any specific stratum of the population even if there is no effect modification or confounding. The admitted mathematical niceties of the OR are not reason enough to accept such a confusing state of affairs. Of course, when the outcome is rare, the OR approximates the RR and is, therefore, approximately collapsible.

The noncollaposibility of OR also brings additional difficulties to its conversion into the RR, RD and the number needed to treat (NNT). Contrary to the ease with which Doi et al suggest that other effect measures can be readily-derived from OR, the simple algebraic conversion noted by Doi et al. converting stratum-specific ORs to stratum-specific RR and RD will not yield the crude RR, RD and NNT one might calculate, for example, by classical Mantel-Haenszel methods.^38,41^

## 3. Is OR “portable” with varying baseline risks in meta-analyses?

Because the values of the aforementioned ORs are highly dependent on the population they are evaluated at, its violation of portability seems inevitable. In fact, effect modification and confounding bias are common in practice such that a direct inference based on estimated measures about external populations, i.e., portability, is hardly satisfied. Doi et al. examined the correlation between ORs and baseline risks in all studies across distinctive meta-analyses and found no correlation. However, they did not examine whether there was a correlation between ORs and baseline risks within individual meta-analyses, which is a more relevant question. We argue that the true dependence of the OR on the baseline risk may be masked by merging miscellaneous meta-analyses. Without considering the differences between meta-analyses, the conclusion in Doi et al., that no correlation between OR with the baseline from a mixture of distinctive meta-analyses implies portability, is concerning.

We suspect that such an assumption may be easily violated if ORs correlate with baseline risks within the *same* meta-analysis. Hence, we assessed the correlation of the ORs with the baseline risks within each meta-analysis using the Cochrane Database of Systematic Reviews (CDSR), which is updated to January 2020. A total of 40,243 meta-analyses containing at least 3 studies with binary outcomes were included. For studies with zero counts where OR is not defined in the corresponding 2×2 table, rather than omitting the studies with the information about rare outcome as in Doi et al., we added 0.5 to all cells of the 2×2 table of the studies with at least one zero count as recommended by a recent study.^42^ The Spearman’s rank correlation coefficient (Spearman’s *ρ*) was used to capture both the linear and nonlinear correlation between the OR and the baseline risk for studies in each meta-analysis. To detect whether such correlation changes with the number of studies per meta-analysis, the quantiles of the Spearman’s *ρ* were summarized, after stratified by number of studies in a meta-analysis. Our summary over CDSR and visualizations were implemented in R (version 3.5.3).

Figure 1 presents the correlation of the OR with the baseline risk with the varying number of studies (≥ 3) per meta-analysis. The OR is noticeably correlated with the baseline risk. In nearly half of 40,243 meta-analyses, the absolute values of Spearman’s *ρ* are larger than 0.5, a threshold corresponding to moderate correlation.^43^ Although the portability appears to hold for OR across varying levels of prevalence from the Figure 3 in Doi et al., our results show that the correlation between the OR and the baseline risk is clearly not negligible in most of the meta-analyses (Figure 1). In fact, those correlations are generally negative; most 75% quantiles of Spearman’s *ρ* are negative across all number of studies.

**Figure 1:**
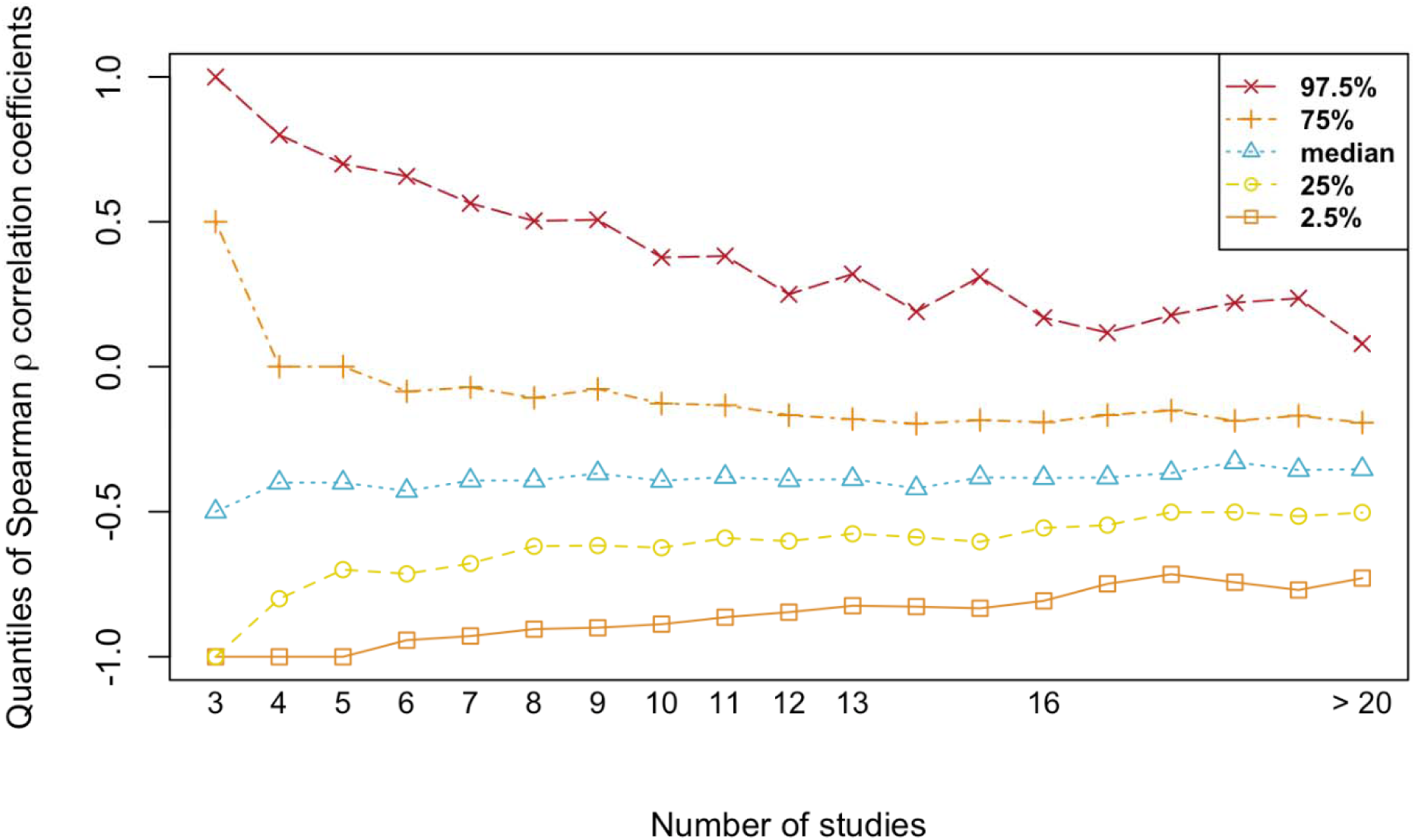
Quantiles of the Spearman’s rank correlation coefficient ρ between the odds ratio and baseline risk among meta-analyses, stratified by number of studies in a meta-analysis.

## 4. What to do next? Estimating overall and conditional effects from a bivariate generalized linear mixed model (BGLMM)

To account for potential correlation of the baseline risk with a treatment (or exposure) effect measure, meta-regression has been widely used to obtain the effects conditioning on the baseline risk in medical literatures.^30,44,45^ However, several pitfalls of a naïve meta-regression have been criticized, because the baseline risk itself is a post-randomization variable rather than a fixed covariate.^46-49^ Alternatively, several studies have recommended using generalized linear mixed-effects models (BGLMMs) to jointly model the effect size and the baseline risk.^33,36,50^

### 4.1 The BGLMM

In this section, we describe the BGLMM model and discuss how to obtain the overall (marginal or population average effect) and the effect conditioning on varying baseline risks to account for potential correlation. To be general, we discuss estimation of both marginal and conditional OR, RR and RD from the BGLMM model. Consider a meta-analysis with *N* studies, BGLMM will jointly model the underlying event risks from the baseline and treatment (or exposure) group.^51^ For the ith study *i* = 1…,*N*, let *p*_*i*0_ denote the baseline risk, and *p*_*i*1_ the risk in the treatment group. As a result of potential correlation of the study-specific treatment effect with the corresponding baseline risk *p*_*i*0_, the underlying true baseline and exposure risks *p*_*i*0_ and *p*_*i*1_ may be correlated. They can be jointly modeled by a BGLMM as:

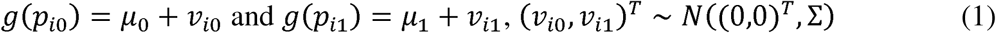

Here, *g*(·) denotes the link function which transforms the event probabilities to linear forms, and 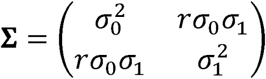 denotes the covariance. Although one can apply other link functions, we choose the most commonly used link function, the logit link. The study-specific treatment effect in the logit scale is therefore expressed as logit(*p*_*i*1_)− logit(*p*_*i*0_), i.e., the log(*OR*_*i*_). The parameters μ_0_ and μ_1_ are fixed effects and represent the average baseline and treatment risks in the logit scale. The corresponding study-specific parameters *v*_*i*0_ and *v*_*i*1_ are random effects and assumed to follow a bivariate normal distribution with covariance **∑**. The parameters 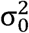 and 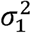 are between-study variances for the baseline and treatment odds, logit(*p*_*i*1_) and logit(*p*_*i*0_). The r is the correlation between the baseline and treatment risks in the logit scale, such that the covariance is *rσ*_0_ *σ*_1_. Hence, the variance of the log(*OR*_*i*_) is 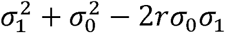 and its correlation with the baseline risk (*p*_*i*0_) in the logit scale *r*_0_ (i.e., baseline log odds) is implied as:

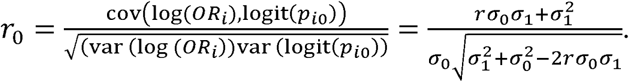

### 4.2 Estimating the marginal effects from the BGLMM

From a BGLMM model with a logit link, the marginal treatment and baseline risks *p*_1_ and *p*_0_ are estimated by 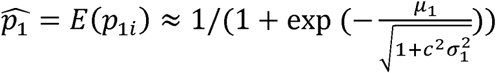 and 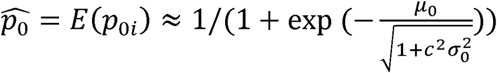 where 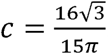^52^. Then, the marginal estimate of OR, RR and RD for any meta-analysis are given 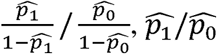 and 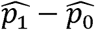.

### 4.3 Estimating the conditional effects from the BGLMM

Similarly, for any given baseline risk *p* in the study *i* (for the prediction purpose can be any integer greater than N in the mixed-effect model), we can obtain the conditional effect estimate based on the conditional mean of the treatment risk *E*(*p*_1*i*_ | *p*_0*i*_ = *p*). According to the bivariate normality assumption between the *v*_*i*0_ and *v*_*i*1_, the conditional mean of treatment risk *p*_1_ in the logit scale is given by:

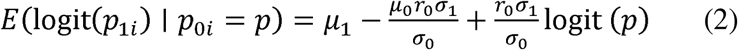

Because the expression to solve for *E*(*p*_1*i*_ |*p*_0*i*_ = *p*) from equation (2) is complex and the median is more robust to skewed distribution than the mean, we follow previous studies and use the median as an estimate of *p*_1*i*_ conditioned on *p*_0*i*_ = *p*.^53^ The conditional median of *p*_1*i*_, i.e., *M*(*p*_1*i*_ |*p*_0*i*_ = *p*), is given by:

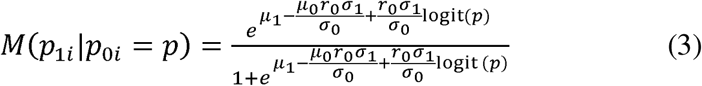

Therefore, the conditional estimate of OR, RD, RR are 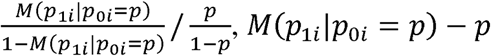 and *M*(*p*_1*i*_ |*p*_0*i*_ = *p*)/*p*, respectively. With all the estimates of fixed-effect parameters, between-study variances, and correlation coefficients from (1), the 95% prediction intervals and confidence intervals (CI) for both the marginal and conditional OR, RD, and RR can be obtained through the delta method and normal approximation.

### 4.4 A case study illustrating the implementation of the BGLMM

As a case study illustrating how to implement BGLMM, we reanalyzed a meta-analysis with 20 studies on comparing the effect of oral sumatriptan (100 mg) versus placebo on mitigating the acute headache at 2 hours after drug administration from the CDSR.^54^ We used PROC NLMIXED in SAS (version 9.4) to fit the BGLMM. The random effects were approximately integrated by the default adaptive Gaussian quadrature, and likelihood maximization used the default dual quasi-Newton optimization algorithm. We also compared the marginal effects estimated from the BGLMM with the overall OR, RR and RD estimated from a standard two-step random-effects meta-analysis without considering correlation as implemented by the rma command in R package “metafor”.^55^ The Spearman’s *ρ* was calculated based on the sample data for each effect summary measure and the corresponding 95% confidence intervals were obtained from Fisher’s *z*-transformation with the variance of *z* estimated by 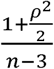.^56^

The Spearman’s *ρ* for all effect measures versus baseline risks are all nonzero. In particular, the Spearman’s *ρ* correlation estimates between the baseline risk and OR, RR and RD are − 0.74 (95% CI, − 0.90 to − 0.39), − 0.90 (95% CI, − 0.97 to − 0.71) and − 0.54 (95%, − 0.81 to − 0.10), respectively. Thus, we fitted the logit BGLMM and Figure 2 shows that the observed OR, RR, RD matched well with the BGLMM. The model obtained an overall (or marginal) OR of 3.48 with 95% CI (2.93 to 4.14), RR (2.05; 95% CI, 1.80 to 2.30) and RD (0.30; 95% CI, 0.26 to 0.33) which are very similar to the OR (3.50; 95% CI, 2.94 to 4.16), RR (2.02; 95% CI, 1.79 to 2.27), and RD (0.29; 95% CI, 0.26 to 0.33) estimated from the two-step random-effect meta-analysis ignoring the correlation.

**Figure 2:**
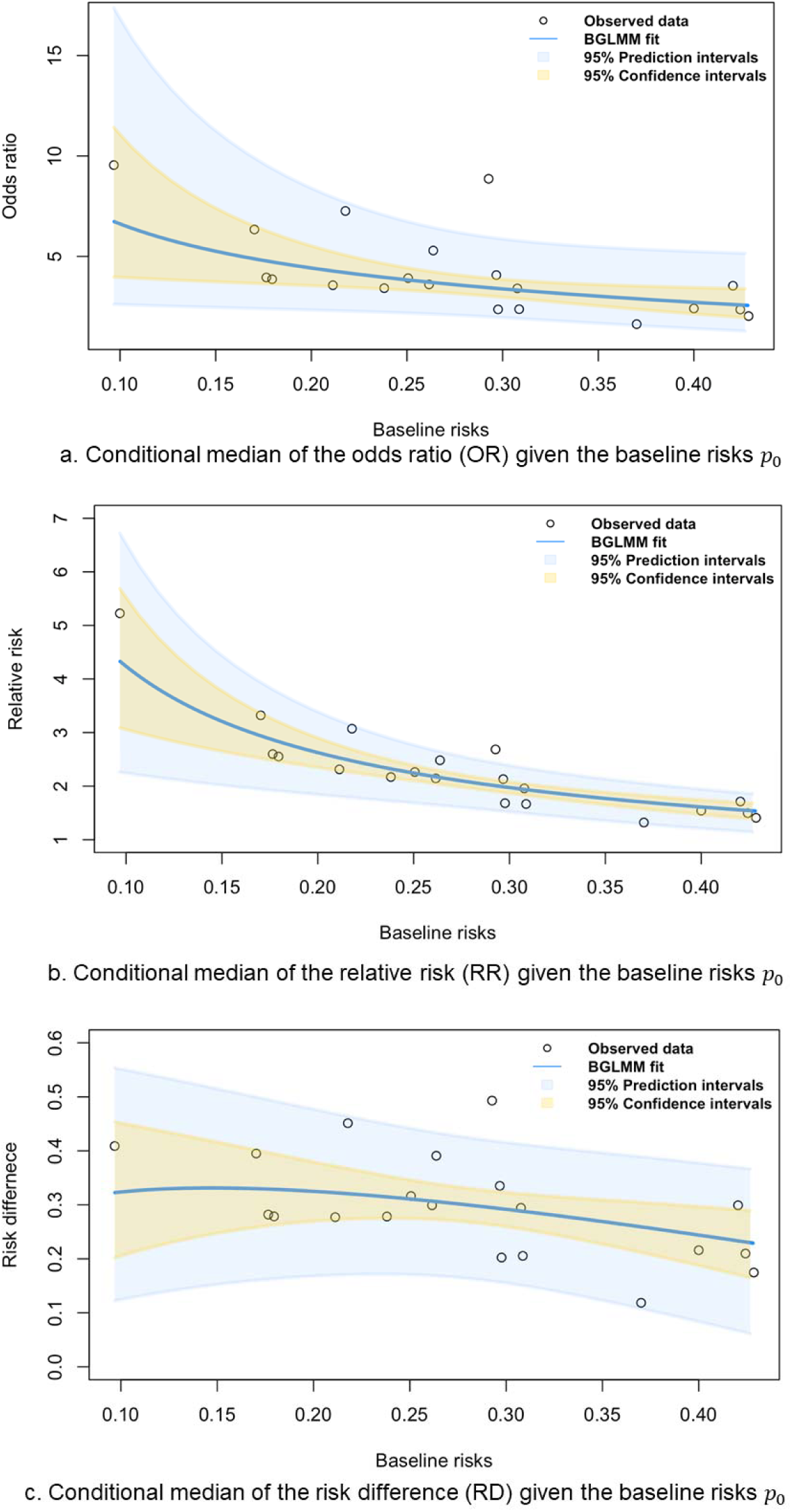
Plot of the observed OR, RR, and RD versus the nonlinear fit from BGLMM with varying levels of baseline risks; BGLMM: bivariate generalized linear mixed-effects model; OR: odds ratio; RD, risk difference; RD, relative risk.

As shown in Figure 2a, the OR is not stable as the baseline risk changes. The OR estimate from the BGLMM decreases from 6.73 to 2.56 when the baseline risk increases from 0.10 to 0.43 (Figure 2a). The conditional RR and RD obtained from the BGLMM show similar trends, although the pattern is less obvious for the conditional RD (Figure 2b and 2c). Furthermore, the estimated correlation 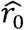 between the log OR and the log baseline odds from the BGLMM is − 0.68 (95% CI, − 0.91 to − 0.14), which strengths the evidence that there is a significant correlation between the treatment effect and the baseline risk. Combined with the fact that most of the observed OR (or RR and RD) as in Figure 2a (or Figure 2b and 2c) are within the 95% prediction interval by BGLMM, it is clear that the correlations between the effect measures and baseline risks are not negligible.

## 5. Discussion

Because of the interpretability issues and lack of collapsibility, we urge researchers to avoid ORs when either the RD or RR is available. Furthermore, through a systematic evaluation of the CDSR and a real-world meta-analysis example, our results suggest that the value of OR depends on the baseline risk when the differences between meta-analyses are considered. Not only may the baseline risk of the outcome be correlated with the corresponding effect measure in the meta-analysis, but it also dictates the meaningfulness of those effect measures for a single study. This dependency underscores the importance to report the baseline measure for a correct effect interpretation. The assumption that OR is likely “portable”, which seems to be the main reason why Doi et al prefer OR over RR, is easily violated when the OR correlates with the risk in the control group within the same meta-analysis. In summary, the recommendation of replacing RR with OR appears misguided.

Even if the OR is not correlated with the baseline risk in a meta-analysis, only relying on a single aspect to replace one measure with the other is near-sighted; other issues such interpretability and collapsibility ought to impact the choice of effect measure. Rather than attempting to set one method aside when the correlation with the baseline risk is of concern, perhaps a more meaningful step is to include the baseline risk and report the variation in the effect measure with baseline risks in addition to the marginal effect, regardless of the measure of choice.

## Data Availability

The data comes from the Cochrane Database of Systematic Reviews.

## Acknowledgements and Funding

The authors gratefully acknowledge the NIH National Library of Medicine (R01LM012982).

